# Evaluation of the Analytical Performance of Hemoglobin Analysis on the Point-of-Care Testing Device HemoCue Hb 801

**DOI:** 10.1101/2025.08.26.25334339

**Authors:** Sophie Vernimmen Olesen, Leif Kofoed Nielsen, Lene Gredal

## Abstract

**Introduction:** Evaluating the accuracy and imprecision of Point-of-Care Testing (POCT) devices is an important prerequisite for ensuring proper diagnosis. The objective of this study was to assess the reliability of POCT hemoglobin (Hb), conducting a method comparison study on HemoCue Hb 801 and Sysmex XN 9000 as reference.

**Methods:** Thirty EDTA whole blood samples previously tested for Hb on Sysmex XN 9000 were also analysed on HemoCue Hb 801 within the measurement range 6.4-21.9 g/dL. Data were assessed by linear regression and correlation with a 95% confidence interval (CI) for the slope and intercept, a paired Student’s t-test, bias calculations and a Bland-Altman plot. Twelve independent measurements on the same sample material determined the imprecision of HemoCue Hb 801.

**Results:** We found a strong correlation coefficient (*r*^*2*^) of 0.992 between Hb measured on HemoCue Hb 801 and Sysmex XN 9000 with a slope of 0.984 (CI: 0.956 to 1.021) and an intercept of 0.181 (CI: -0.283 to 0.544). The paired Student’s t-test indicated that the mean Hb differences between the methods were not statistically significant (p=0.84). Bias (-0.01 g/dL) did not deviate from the allowed maximum absolute bias (±0.35 g/dL). The values were within the 95% limits of agreement (±1.40 g/dL) for the differences. The coefficient of variation was 1.0%.

**Conclusion:** There was a strong correlation and good agreement between Hb measured on HemoCue Hb 801 and Sysmex XN 9000, as well as an acceptable bias and imprecision, supporting a good accuracy.

## 1. Introduction

The use of near-patient testing or Point-of-Care Testing (POCT) is growing rapidly in hospitals, including emergency departments, but is also used in medical practice, outpatient clinics, nursing homes and in patients’ homes. POCT devices determine, among others, a diversity of hematological biomarkers, e.g. Hb, being a relevant near-patient analysis. POCT Hb devices can provide early and rapid results by reducing transport time and expanding laboratory testing capacity. Therefore, POCT Hb devices have an advantage in anemia screening, therapy control, and in situations where analytical results initiate decisions regarding acute treatment options for life-threatening anemic conditions. Furthermore, POCT Hb devices can be used as a supplement to the automated hematology analyses in the clinical biochemical departments and in general strengthen the link between prevention, diagnostics, and treatment in the healthcare system.^1–3^

Hemoglobin concentrations analysed on the POCT device HemoCue Hb 801 (HemoCue) are determined by the absorbance of whole blood measured at an Hb/HbO_2_ isosbestic point, a wavelength where two different absorption spectra intersect. The absorbance of the sample, thus the Hb concentration, will be independent of whether Hb is oxidized or not, i.e. independent of the saturation. The measuring wavelengths are 506 and 880 nm, the latter used to compensate for lipemia, bilirubin and leucocytes, which affect turbidity, causing false elevated Hb concentrations. The HemoCue Hb 801 System is calibrated against the international reference method for Hb determination, the hemiglobincyanide method.^1,4^

The Department of Clinical Biochemistry, Hvidovre Hospital (DCB), is responsible for the quality assurance of POCT devices used outside the central laboratory. Valid POCT Hb measurements are an important prerequisite for ensuring that patients are diagnosed properly and receive proper treatment. It is therefore clinically imperative to examine whether the HemoCue Hb 801 System complies with certain quality-specific requirements before this POCT equipment is put into service.^1^

This study aimed to evaluate the performance of Hb measurements on the HemoCue Hb 801 System by performing a method comparison between HemoCue Hb 801 and the reference method Sysmex XN 9000 (Sysmex) on EDTA-stabilized whole blood. Also, we wanted to determine the degree of agreement between independent Hb measurements on the HemoCue Hb 801 System.

## 2. Materials and Methods

### 2.1 Specimens

Venous whole blood was collected in 4 mL K_2_EDTA vacuette tubes (Greiner Bio-One) according to manufacturer’s instructions. The samples were kept at room temperature until analysis.^4–6^ Analyses were performed within 24 hours on Sysmex XN 9000 and HemoCue Hb 801.

Thirty patient samples with evenly distributed Hb concentrations throughout the defined measurement range of HemoCue Hb 801 (6.4-21.9g/dL)^4^ were selected based on previous analysis on Sysmex XN 9000. Ten samples were collected within each of three ranges: [6.4 – 9.7] g/dL; [9.8 – 14.4] g/dL, and [14.5 – 21.9] g/dL.

The study complied with all the relevant national regulations and institutional policies and is in accordance with the tenets of the Helsinki Declaration. According to the Danish National Committee on Health Research Ethics, ethical approval was not required, since the samples were anonymized, no additional samples were collected due to the study’s retrospective design, and the measurements were used solely for method comparison.

### 2.2 Analytical Methods

According to HemoCue Hb 801 instructions, sample tubes were mixed manually by reversing the tubes 10 times before each measurement. Diff-safe blood dispensers (Alpha Scientific Corporation) were applied to the test tubes, and a drop of sample material was placed on parafilm (Amcor), from which the HemoCue Hb 801 microcuvettes were filled for measurement on the HemoCue Hb 801 Analyzer.^4^

To examine the imprecision of HemoCue Hb 801, 12 independent measurements were carried out on one sample. The sample selected had a Hb concentration of 14.0 g/dL measured on Sysmex XN 9000.

The quality control procedures for Sysmex measurements were followed, and the Hb controls were within the acceptable limits. The HemoCue Hb 801 Analyzer has a builtin quality control, which, at each start-up and at every hour when the device is still used, automatically checks the optics.^4^

### 2.3 Data Analysis

Statistical analyses were performed using Microsoft Excel 2016 (Microsoft Corporation) and IBM SPSS Statistics for Windows, Version 28.0.1.1 (IBM Corp), and Microsoft Excel 2016 to generate graphs. Data were evaluated for Gaussian distribution using histograms, Q-Q plots, skewness, kurtosis and by use of Shapiro-Wilks test for normality.

Correlation and agreement between Hb concentrations measured on HemoCue Hb 801 and Sysmex XN 9000 were assessed by linear regression. The correlation was determined by using the correlation coefficient r^2^, and the 95% confidence interval (CI) for the slope and intercept was estimated.

The Student’s paired t-test was used to evaluate the statistical significance of the difference between the mean values of the two methods. A p-value of < 0.05 was considered significant.

DCB is accredited by the Danish Accreditation Fund (DANAK) under ISO/IEC 15189^7^ and has set the maximum allowed absolute bias at 10% of the difference between the upper and lower limits of the local reference range (men: [13.4–16.9] g/dL; women: [11.8–15.3] g/dL), corresponding to ±0.35 g/dL. Bias calculations were performed to assess whether the mean bias exceeded this locally defined threshold.

Furthermore, the 95% limits of agreement (LoA) were used to assess if the differences in Hb values were of clinical importance. The 95% LoA were defined by DCB as 4 times the maximum absolute bias (i.e., ±1.40 g/dL).

In addition, a Bland-Altman^8^ plot was used to evaluate and visualize the degree of agreement (bias) between Hb measured on HemoCue Hb 801 and Sysmex XN 9000.

Determination of the imprecision of HemoCue Hb 801 was performed to evaluate the accuracy of the POCT device. To examine the imprecision, mean, standard deviation, and coefficient of variation (CV) were calculated from 12 independent Hb measurements on one sample and compared with the stated CV by the manufacturer (HemoCue) and the locally defined CV for Sysmex.

## 3. Results

Linear regression analysis shown in Figure 1 produced a correlation coefficient (*r*^*2*^) of 0.992, a slope of 0.984 (CI: 0.956 to 1.021) and an intercept of 0.181 (CI: -0.283 to 0.544).

**Figure 1.**
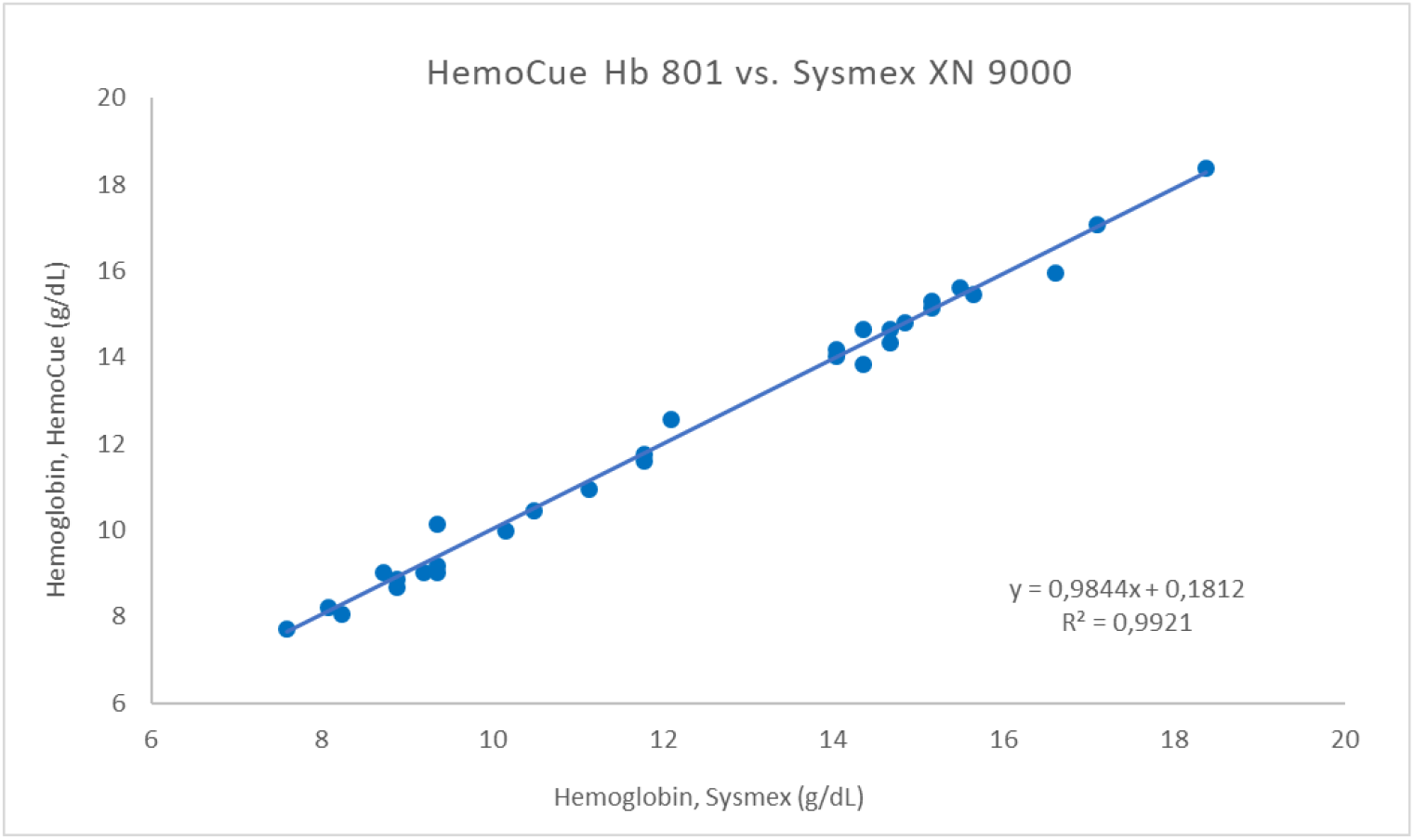
Linear regression and correlation for hemoglobin measured on HemoCue Hb 801 compared with Sysmex XN 9000. The x-axis represents the hemoglobin value in g/dL measured by Sysmex, and the y-axis the hemoglobin value in g/dL measured by HemoCue Hb 801.

No significant difference was found between HemoCue Hb 801 and Sysmex XN 9000 (P=0.84).

The mean bias between HemoCue Hb 801 and Sysmex XN 9000 was -0.01 g/dL, which did not deviate from the locally allowed maximum absolute bias of ± 0.35 g/dL.

A Bland-Altman plot is depicted in Figure 2, visualizing the agreement between Hb concentrations analysed on Sysmex XN 9000 and HemoCue Hb 801. The x-axis indicates the mean of the measured Hb values from the two methods, and the y-axis denotes the difference between the two measured values (HemoCue Hb 801 - Sysmex XN 9000). The mean bias (-0.01 g/dL), the allowed maximum absolute bias (±0.35 g/dL) and the 95% LoA (±1.40 g/dL) are also depicted.

**Figure 2.**
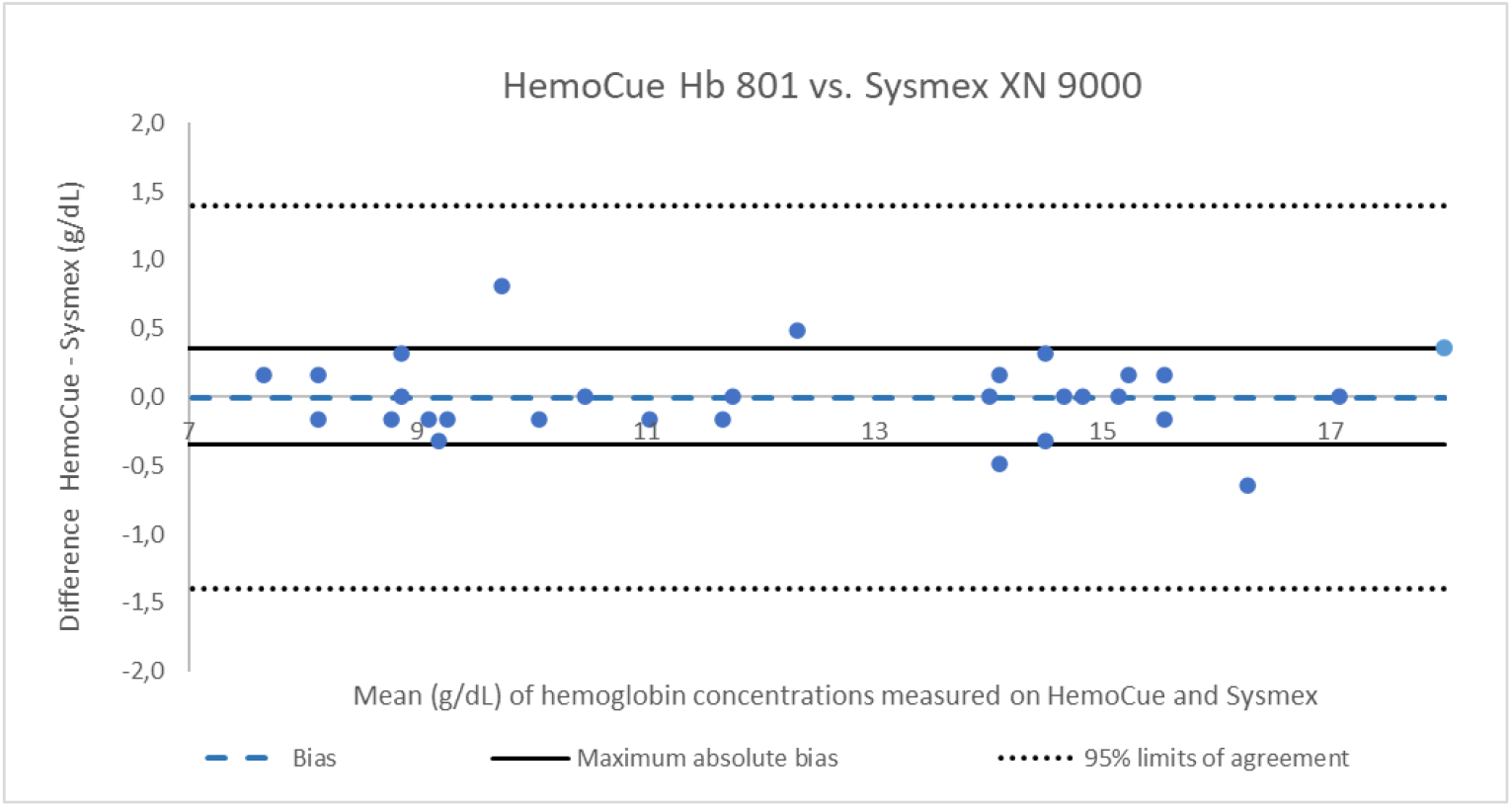
Bland-Altman Plot for hemoglobin concentrations measured on HemoCue Hb 801 and Sysmex XN 9000. The mean values of the hemoglobin measurements for the two methods are presented on the x-axis and the differences on the y-axis. The blue-dotted line represents the mean bias of -0.01 g/dL. The black lines define the upper and lower limits for the maximum absolute bias (± 0.35 g/dL). The black-dotted lines refer to the upper and lower 95% limits of agreement (± 1.40 g/dL) for the differences.

The calculated CV for HemoCue Hb 801 was 1.0%, which did not exceed the CV stated by the manufacturer of 1.0%.

## 4. Discussion

The present investigation aimed to compare Hb concentrations measured on HemoCue Hb 801 System with Sysmex XN 9000 as the reference method, evaluating the reliability of the analytical performance of the POCT device.

The linear regression analysis with a correlation coefficient (*r*^*2*^) of 0.992 showed a strong correlation between the two methods, as well as a good agreement, since the intercept with the y-axis (0.181) was close to 0 and the slope (0.984) was close to 1. The 95% CI around the slope and intercept contained the values 1 and 0, and therefore, not significantly different from the identity line, which indicates that the analytical performance of HemoCue Hb 801 corresponds well to the reference method Sysmex XN 9000.

The good agreement between the two methods was supported by a paired Student’s t-test, showing no significant difference between the two methods (P = 0.84) as well as the Bland-Altman plot, where the values were evenly distributed around zero throughout the whole measurement range. The mean bias between HemoCue Hb 801 and Sysmex XN 9000 was -0.01 g/dL and did therefore not deviate from the allowed maximum absolute bias of ± 0.35 g/dL. The largest difference of 0.81 g/dL did not exceed the 95% LoA of ±1.40 g/dL, which complied with the defined requirements by DCB.

The CV for HemoCue Hb 801 (1.0%) was identical to the stated CV for control level 3 (15.36 g/dL) from the manufacturer (HemoCue)^4^. It was expected that the CV would be lower than the stated CV of 1.0%, since this experimental imprecision evaluation did not include all potential analytical variations, such as performing measurements on different days and on different HemoCue Hb 801 Analysers. Only one measurement (13.9 g/dL) in the imprecision evaluation differed from the other 11 measurements, which all gave the same analytical result of 14.3 g/dL. The stated CV (1.0%) by HemoCue Hb 801 was lower compared with the locally defined CV for Sysmex XN 9000 (2.5%) at DCB. However, the CV value for Sysmex XN 9000 is calculated based on an average from 3 devices and therefore contains several sources of analytical variation.

The POCT equipment HemoCue Hb 801 is intended for use on both capillary and venous blood. However, capillary blood was not chosen since this would include variations of subjective matters from the clinical staff collecting the specimens from finger sticks. The capillary measurements would therefore not exclusively reflect the HemoCue Hb 801 Analyser’s ability to produce valid and precise Hb concentration results.

Also, under normal circumstances, the HemoCue Hb 801 device is validated with controls. However, the lack hereof was not considered problematic, because Sysmex XN 9000 was used as reference method, which at DCB is accredited according to DANAK ISO/IEC standard 15189.^7^ The analytical results from Sysmex XN 9000 therefore represent the true value of Hb concentrations and the method comparison serves the purpose of validating the accuracy of HemoCue Hb 801 itself.

Overall, we found good agreement between Hb measured on HemoCue Hb 801 and Sysmex XN 9000, with a strong correlation. In addition, the performance of the imprecision did not exceed the limits stated by HemoCue, and thus HemoCue Hb 801 is considered reliable for measuring Hb concentrations.

Our findings correspond very well with those from *Young et al*., showing a strong correlation (*r*^*2*^ = 0.88) between Hb measured on Hb 801 and the golden standard (Sysmex XN9100) with a bias of 0.2 g/dL.^2^ Therefore, HemoCue Hb 801 can be implemented as a supplement to Sysmex in the biochemistry laboratories, emergency departments, medical practice, institutions etc.

It should be noted that the analytical results obtained from the HemoCue Hb 801 System are mainly used for anemia screening and therapy control, where in-depth analyses on Sysmex and other analytical equipment in hospital laboratories are used to determine the etiology based on erythrocyte parameters such as red blood cell count (RBC), middle cell volume (MCV), middle cell hemoglobin (MCH), middle cell hemoglobin concentration (MCHC), and other supporting biomarkers e.g. reticulocytes and the plasma components transferrin, ferritin, bilirubin, haptoglobin, vitamin B_12_ and iron. Based on an overall assessment of the individual biomarkers and erythrocyte parameters, a correct diagnosis can be made, thus ensuring the right treatment for the patient.^9^

Finally, training and guidance on the use of the equipment for health professionals, including the education on pre-analytical factors, internal and external quality assurance, analytical methods, maintenance, etc., are factors of great importance for the quality of analytical results of Hb measured on the HemoCue Hb 801.^10^

## Data Availability

All data produced in the present study are available upon reasonable request to the authors

## Patient consent for publication

Not applicable.

## Funding and Acknowledgements

We thank HemoCue for supplying the equipment and microcuvettes for the study. Particularly, we are grateful for the advice and critical review of the manuscript provided by Henriette Lorenzen, Associate Professor at the Faculty of Health, Copenhagen University College.

## Author Contributions

SVO and LG conceived of and designed the study. SVO collected samples. SVO ran experiments. SVO, LG and LKN performed data analysis and interpreted the data. SVO prepared the manuscript draft. SVO, LG and LKN revised the manuscript for important intellectual content, provided final manuscript approval, and agreed to be accountable for all aspects of the work.

## Data availability statement

Data is available on request from the authors.

